# Intensive Blood Pressure Lowering Improves Left Ventricular Geometry in Older Hypertensive Patients: The STEP Trial

**DOI:** 10.1101/2024.03.04.24303756

**Authors:** Yue Deng, Juyan Zhang, Zhenzhen Chen, Jingjing Bai, Xiaomin Yang, Chunli Yu, Jing Yu, Jun Cai, STEP Study Group

## Abstract

**Background:** The effect of intensive systolic blood pressure (SBP) lowering on left ventricular (LV) geometry remains unclear.

**Methods:** Patients with hypertension aged 60–80 years without prior stroke were enrolled from 42 centers across China from January 2017. Eligible patients were randomly assigned to intensive (target: 110 to <130 mmHg) or standard (target: 130 to < 150 mmHg) SBP-lowering treatment. LV mass (LVM) was assessed by two-dimensional, M-mode, color flow Doppler echocardiography. LV hypertrophy (LVH) was diagnosed according to LVM index (LVMI) to height^1.7^ using sex-specific thresholds.

**Results:** Baseline demographic and LV parameters were comparable between the groups (n = 5709). During the median follow-up of 2.63 years, intensive treatment was associated with a lower risk of new LVH (hazard ratio [HR] 0.83, 95% confidence interval [CI] 0.69–1.00, *P* = 0.051) and greater regression of mean LVMI by 0.38 g/m^1.7^ per year (95% CI 0.05–0.71, *P* = 0.024). The rate of baseline LVH regression did not differ between the groups. Male patients achieved a significantly greater benefit from intensive treatment than female patients in terms of new LVH prevention and baseline LVH regression (both *P* for interaction <0.10). The favorable effect of intensive treatment on the cardiovascular outcome (HR 0.73, 95% CI 0.54–1.00) was slightly attenuated after adjusting for LVMI as a time-varying covariate (HR 0.75, 95% CI 0.55–1.03).

**Conclusions:** In older patients with hypertension, intensive SBP lowering offers additional cardiovascular benefits in terms of LV geometry. This favorable effect partially explains the reduction in cardiovascular events.

**Trial registration:** Clinicaltrials.gov (Identifier: NCT03015311)

**Condensed Abstract:** This is the first randomized controlled trial with a sufficient sample size to compare the effect of intensive systolic blood pressure (SBP) lowering (<130 mmHg) on echocardiographic left ventricular (LV) geometry with that of standard SBP lowering (< 150 mmHg) in older patients with hypertension. A lower risk of new LV hypertrophy development and greater regression of LV mass index were observed in patients with intensive treatment than in those with standard treatment. This favorable effect partially explains the reduction in cardiovascular events associated with intensive SBP lowering.

## INTRODUCTION

The left ventricle is the primary end-organ target of hypertension, and left ventricular (LV) remodeling is common in patients with hypertension as a result of chronic blood pressure (BP) overload(1). LV geometric remodeling, particularly the development of LV hypertrophy (LVH), is a proven biomarker of subclinical cardiac damage and a major prelude of incident cardiovascular events, including acute coronary syndrome, stroke, heart failure, and sudden death(2, 3). Successful BP reduction has been demonstrated to prevent the development of new LVH and to promote the regression of existing LVH, in turn improving cardiovascular outcomes(4–7).

Emerging evidence shows that LVH develops in a number of patients at BP values below the range for hypertension as currently defined, such as patients with high-normal BP (systolic BP [SBP] 120–139 mmHg and/or diastolic BP 80–89 mmHg)(8–10) and in patients with hypertension receiving standard BP control therapy(11). Large-sample randomized controlled trials (RCTs) have shown that intensive SBP-lowering therapy (SBP target <120 mmHg) versus standard SBP-lowering therapy <140 mmHg is associated with a lower risk of new LVH on electrocardiography (ECG) in patients with hypertension with a high cardiovascular risk, regardless of diabetes mellitus(12, 13). Similar results have been reported for patients aged ≥55 years in a study comparing SBP lowering to <130 mmHg versus to <140 mmHg(14). Consistently, new ECG-LVH develops less frequently in 60– 80-year-old patients with hypertension with intensive SBP lowering to <130 mmHg than in those with an SBP target of <150 mmHg(15).

Of note, ECG is a low-sensitivity (20%–50%) technique for LVH diagnosis, that ECG-LVH may be influenced by factors other than anatomic LV mass (LVM), such as increased myocardial tension and neurohormonal or biochemical changes(16–18).On the contrary, the SPRINT-HEART study observed no difference in LV mass changes detected by cardiac magnetic resonance imaging in patients who received intensive SBP lowering compared with those who received standard SBP lowering(19). Thus far, few large-sample RCTs have provided direct evidence on the favorable effect of intensive SBP lowering on imaging LV geometry. Additionally, despite observational studies indicating a clear association of LVH with adverse outcomes, recent RCTs that have demonstrated the benefits of intensive SBP lowering on LV geometry could not explain most of the reduction in cardiovascular events associated with BP lowering(12, 13, 15). The role of LV geometry in mediating hypertension-related cardiovascular events remains inconclusive.

In the Strategy of Blood Pressure Intervention in the Elderly Hypertensive Patients (STEP) trial, older patients with hypertension were randomly assigned to intensive SBP-lowering therapy (SBP target: 110 to <130 mmHg) or standard SBP-lowering therapy (130 to <150 mmHg), and the clinical cardiovascular prognosis was compared between the two groups(20, 21). Each patient underwent standard echocardiography at baseline and at follow-up. This trial provided a unique opportunity to examine the effects of intensive SBP lowering on LV geometry and its mediation role in the reduction of cardiovascular risk associated with intensive SBP lowering in older patients with hypertension.

## METHODS

### Study population and design

The data that support the findings of this study are available from the corresponding author upon reasonable request. The STEP trial was a prospective, multicenter, RCT that included 42 clinical centers throughout China. In the study, the clinical outcomes of older patients with hypertension who underwent intensive SBP lowering (SBP target: 110 to <130 mmHg) versus standard SBP lowering (SBP target: 130 to <150 mmHg) were evaluated. All patients were aged 60–80 years, of Han ethnicity, and had an SBP of 140–190 mmHg or were on antihypertensive therapy. The exclusion criteria were previous stroke, mental impairment, uncontrolled diabetes mellitus, or a serious life-limiting condition. The design and results of the STEP trial have been reported previously(20, 21). This trial was approved by the ethics committee of FuWai Hospital and all collaborating centers. The trial was conducted in accordance with the principles outlined in the Declaration of Helsinki. All enrolled patients provided written informed consent.

### Intervention, BP measurements, and follow-up

All enrolled patients were treated with antihypertensive medications, including olmesartan, amlodipine, and if needed, hydrochlorothiazide. Office BP was measured by trained trial staff members using the same validated office BP monitor (Omron Healthcare Group, Kyoto, Japan). Sitting brachial BP was measured at the upper right arm after 5 min of rest using an appropriately sized cuff, and the average of three readings obtained at 1-minute intervals was calculated.

Patients were scheduled for follow-up visits monthly during the first 3 months and every 3 months thereafter. At each visit, a structured interview was performed in both treatment groups to obtain self-reported events. The primary outcome was defined as a composite of stroke, acute coronary syndrome, acute decompensated heart failure, atrial fibrillation, and cardiovascular death. All events were evaluated by an independent endpoint adjudication committee blinded to treatment allocation.

### Echocardiography measurements

Standard two-dimensional, M-mode, pulsed, continuous wave, color flow Doppler echocardiography was performed for each subject at baseline and at the follow-up. Echocardiographic raters across all sites were trained to obtain the measurements of LV parameters and other cardiac parameters in accordance with the recommendations of the American Society of Echocardiography (ASE)(22). The LV end-systolic internal diameter (LVIDs), LV end-diastolic internal diameter (LVIDd), interventricular septal end-diastolic thickness (IVSTd), and posterior wall end-diastolic thickness (PWTd) were measured in the parasternal long-axis view at the level of the mitral valve leaflet tips. LV ejection fraction (LVEF) was measured in the apical two-chamber view using the biplane Simpson method. LVM was calculated using the Devereux formula, as follows: LVM (g) = 0.8 × 1.04 ([IVSTd + PWTd + LVIDd]^3^ − [LVIDd]^3^) + 0.6(23). Relative wall thickness (RWT) was calculated as RWT = (PWTd × 2) ÷ LVIDd(24).

The LVM was indexed to height^1.7^ (H^1.7^) as proposed by Chirinos et al(25). LVH was diagnosed if LVM/H^1.7^ was >81 g/m^1.7^ in males or >78 g/m^2^ in females(26). In the sensitivity analysis, LVM was indexed to H^2.7^ based on the ASE recommendations(27), LVH was diagnosed when LVM/H^2.7^ was >48 g/H^2.7^ in males and >49 g/H^2.7^ in females.

### Intra-rater and inter-rater variability

To test the intra-rater and inter-rater variability of the echocardiographic measurements, three key parameters, including IVSTd, LVIDd, and PWTd, were remeasured in 40 randomly selected patients (20 males and 20 females) in the analysis. Intra-rater variability was assessed by Z.J., and inter-rater variability was assessed by two investigators (Z.J. and Y.X.).

### Statistical analysis

Continuous variables were compared between the groups using the *t*-test or Wilcoxon’s rank-sum test. Categorical variables were compared using the likelihood ratio chi-square test or Fisher’s exact test. Multivariate imputation was used for missing data.

To compare the time to detection of LVH in patients without baseline LVH and the time to regression of LVH in patients with baseline LVH, the Cox proportional hazards regression model was used with stratification by clinical site and adjustment for potential covariates (including age, sex, body mass index [BMI], baseline SBP, baseline total cholesterol, baseline estimated glomerular filtration rate, smoking status, diabetes mellitus, and chronic heart disease). The follow-up time was censored on the date of the last echocardiography.

Interactions between the treatment effect and the pre-specified subgroups, namely age (<70 years vs. ≥70 years), sex (male vs. female), SBP tertile (≤138 vs. >139 to <151 vs. ≥152 mmHg), and diabetes mellitus status (yes vs. no), were assessed using a likelihood ratio test for interaction. To examine whether the impact of intensive BP-lowering treatment on the primary outcome could be explained by its impact on LVH, we examined the magnitude of attenuation in the association between intensive SBP lowering and the primary outcome after adjusting for LVH or LVMI as time-varying covariates. The Bland–Altman analysis was used to assess intra-rater and inter-rater variability.

All statistical analyses were performed using R software, version 3.6.3 (R Foundation for Statistical Computing, Vienna, Austria). A two-sided *P* value of <0.05 was considered statistically significant.

## RESULTS

### Baseline demographic and echocardiographic characteristics

After excluding patients with missing or uninterpretable echocardiography results at baseline (n = 1367) or at follow-up (n = 1435), a total of 5709 patients from the STEP trial were included in the analysis. Of these, 2915 were randomly assigned to intensive treatment and 2794 to standard treatment (Figure S1). The baseline demographic characteristics of the enrolled patients were well-matched between the treatment groups, with the mean age being 66.1 years, 46.3% being male, and 18.8% having diabetes mellitus. The exception was that BMI was lower in patients with intensive treatment than in patients with standard treatment (25.6 vs. 25.7 g/m^2^, respectively, *P* = 0.025) (Table 1). The demographic characteristics of the enrolled patients (versus excluded patients) are shown in Table S1. Collectively, the excluded patients were older, had lower baseline BP and BMI values, and had a lower prevalence of chronic kidney disease.

**Table 1.** Baseline demographic and clinical characteristics of the patients.

| Characteristics | Intensive treatment<br>(N = 2915) | Standard treatment<br>(N = 2794) | P-value |
| --- | --- | --- | --- |
| Age, years | 66.1 ± 4.8 | 66.1 ± 4.7 | 0.832 |
| Age ≥70 yr, no. (%) | 674 (23.1) | 632 (22.6) | 0.675 |
| Male, no. (%) | 1348 (46.2) | 1298 (46.5) | 0.893 |
| **Body mass index, kg/m<sup>2</sup>** | **25.6 ± 3.1** | **25.7 ± 3.2** | **0.025** |
| Baseline blood pressure, mmHg |  |  |  |
| Systolic | 146.4 ± 17.0 | 146.4 ± 16.7 | 0.989 |
| Diastolic | 82.8 ± 10.7 | 82.5 ± 10.7 | 0.378 |
| Distribution of SBP, no. (%)<sup>*</sup> |  |  | 0.906 |
| ≤138 mmHg | 926 (31.8) | 901 (32.2) |  |
| 139-151 mmHg | 965 (33.1) | 912 (32.6) |  |
| ≥152 mmHg | 1024 (35.1) | 981 (35.1) |  |
| Fasting serum glucose, mmol/L | 6.1 ± 1.6 | 6.1 ± 1.6 | 0.436 |
| eGFR < 60 ml/min/1.73m<sup>2</sup>, no. (%) | 41 (1.4) | 43 (1.5) | 0.429 |
| Lipid profile, mmol/L |  |  |  |
| Total cholesterol | 4.9 ± 1.1 | 4.9 ± 1.1 | 0.480 |
| Triglycerides (IQR) | 1.3 (1.0, 2.0) | 1.3 (1.0, 1.9) | 0.570 |
| High-density lipoprotein cholesterol | 1.3 ± 0.3 | 1.3 ± 0.3 | 0.422 |
| Low-density lipoprotein cholesterol | 2.7 ± 0.9 | 2.7 ± 0.9 | 0.586 |
| Smoking, no. (%) | 465 (16.0) | 456 (16.3) | 0.732 |
| Medical history, no. (%) |  |  |  |
| Diabetes mellitus | 544 (18.7) | 529 (18.9) | 0.819 |
| Hyperlipidemia | 1076 (36.9) | 1040 (37.2) | 0.830 |
| Cardiovascular diseases | 167 (5.7) | 176 (6.3) | 0.395 |
| The 10-year Framingham risk score<br>≥15%<sup>†</sup>, no. (%) | 2123/2748 (77.3) | 2010/2618 (76.8) | 0.700 |

Values are presented as the mean ± standard deviation, the median (interquartile range), or number (percentage), as appropriate. The P-value were calculated using the t-test or Wilcoxon's rank-sum test. <sup>\*</sup>The distribution of systolic blood pressure is presented as the tertile at baseline. <sup>†</sup>A 10-year Framingham risk score of ≥15% indicates high cardiovascular risk. eGFR = estimated glomerular filtration rate, IQR = interquartile range, SBP = systolic blood pressure. N = 2915 in the intensive treatment group, N = 2794 in the intensive treatment group.

In Table S2, the sex-specific values for baseline LV parameters, including IVSTd, LVIDs, LVIDd, PWTd, and LVEF, were comparable between the treatment groups. The baseline LVM/H^1.7^ was comparable between the intensive-treatment group and the standard-treatment group (70.4 vs. 70.5 g/m^1.7^ in males; 68.2 vs. 68.4 g/m^1.7^ in females). Consistently, the prevalence of baseline LVH was similar between the treatment groups (22.9% vs. 22.4% in males; 23.4% vs. 24.0% in females). These results were essentially the same when LVM was indexed to H^2.7^. Other baseline echocardiographic parameters, including the diameter of the aortic sinus, left atrium, pulmonary artery, and right ventricular outflow tract; end-diastolic volume; prevalence of LVEF <50%; and prevalence of E/A <1, were all comparable between the groups.

### BP and LVH throughout follow-up

Throughout the median follow-up of 2.63 years, the mean BP was 126.8/76.4 mmHg in the intensive treatment group and 136.4/79.3 mmHg in the standard treatment group, resulting in a between-group difference of 9.6/2.9 mmHg (both *P* < 0.001) (Figures S2 and S3). The between-group BP difference were comparable between sex (9.4/2.8 mmHg in females vs. 9.7/3.0 mmHg in males).

Among the patients in the STEP trial without LVH at baseline (n = 4383), the number of cases of newly developed LVH was marginally lower in the intensive treatment group (n = 214, 9.6%) than in the standard treatment group (n = 260, 12.1%) (hazard ratio [HR] 0.83, 95% confidence interval [CI] 0.69–1.00, *P* = 0.051). This beneficial effect was more pronounced in males (HR 0.72, 95% CI 0.54–0.95, *P* = 0.020) than in females (HR 0.94, 95% CI 0.74–1.22, *P* = 0.688) (*P* for interaction = 0.015). The protective effect of intensive SBP lowering against new LVH development was also evident in patients with the highest SBP tertiles (HR 0.71, 95% CI 0.52–0.96, *P* = 0.027), and was consistent across the prespecified subgroups for age, SBP tertile, and diabetes mellitus status (all *P* for interaction > 0.10) (Figure 1).

**Figure 1.**
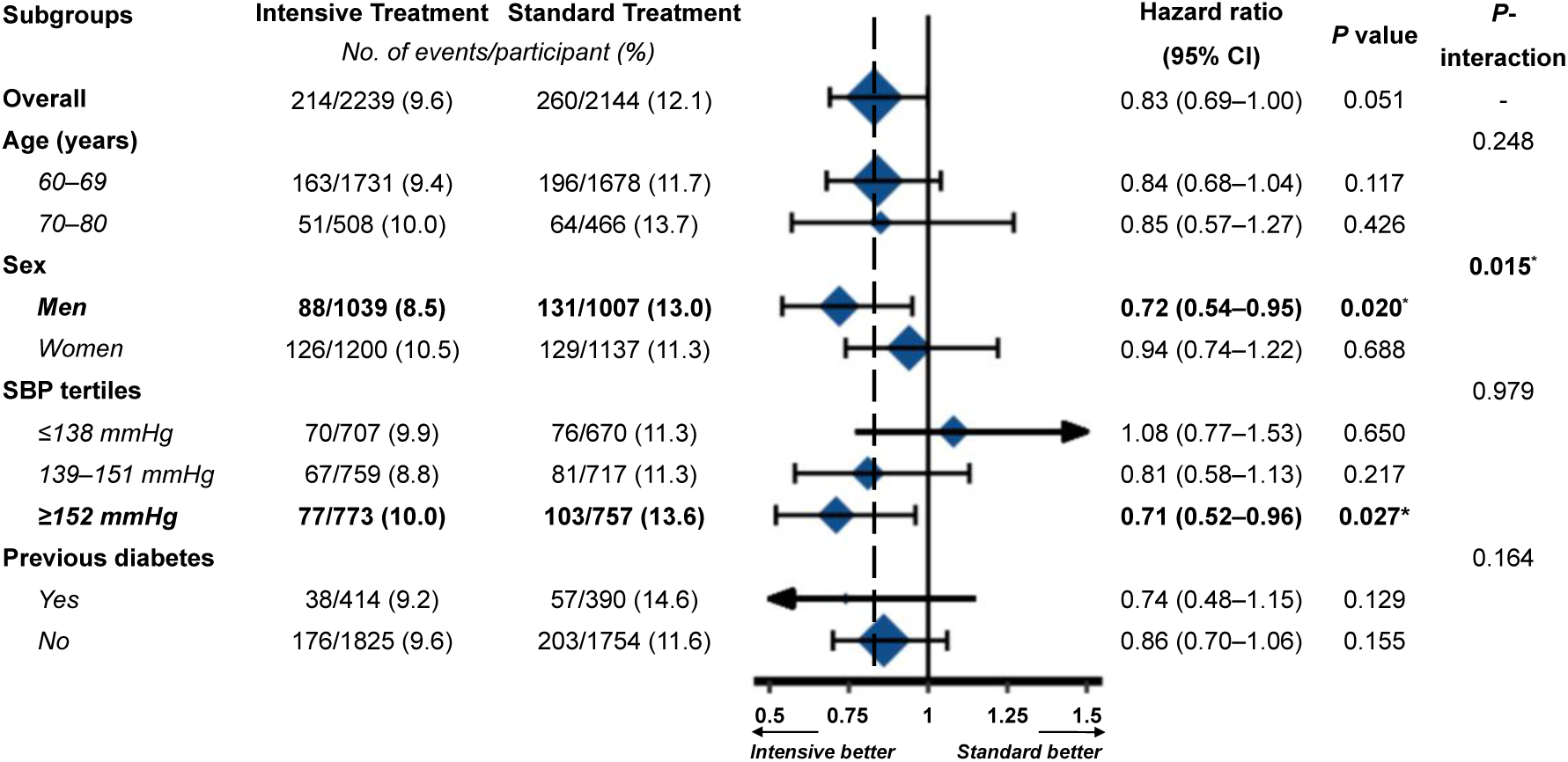
Forest plot showing the effect of intensive (versus standard) SBP-lowering treatment on LVH development among the pre-specified subgroups during follow-up. LVH was diagnosed based on LVM/H^1.7^. The results are presented as the HR and 95% CI. The dashed line represents the overall HR. The *P*-value was calculated using Cox proportional hazards regression model. CI = confidence interval, HR = hazard ratio, H^1.7^ = height^1.7^, LVH = left ventricular hypertrophy, SBP = systolic blood pressure. N = 2239 in the intensive treatment group, N = 2144 in the intensive treatment group.

Among the patients in the STEP trial with LVH at baseline (n = 1326), the rate of LVH regression was comparable between the treatment groups (HR 1.10, 95% CI 0.95–1.28, *P* = 0.221). Notably, intensive (versus standard) SBP lowering was significantly associated with LVH regression in males (HR 1.32, 95% CI 1.04–1.68, *P* = 0.025), but not in females (HR 0.99, 95% CI 0.81–1.22, *P* = 0.924) (*P* for interaction = 0.049). The effect of intensive SBP lowering on baseline LVH regression was consistent among the subgroups for age, SBP tertiles, and diabetes mellitus status (all *P* for interaction > 0.10) (Figure 2).

**Figure 2.**
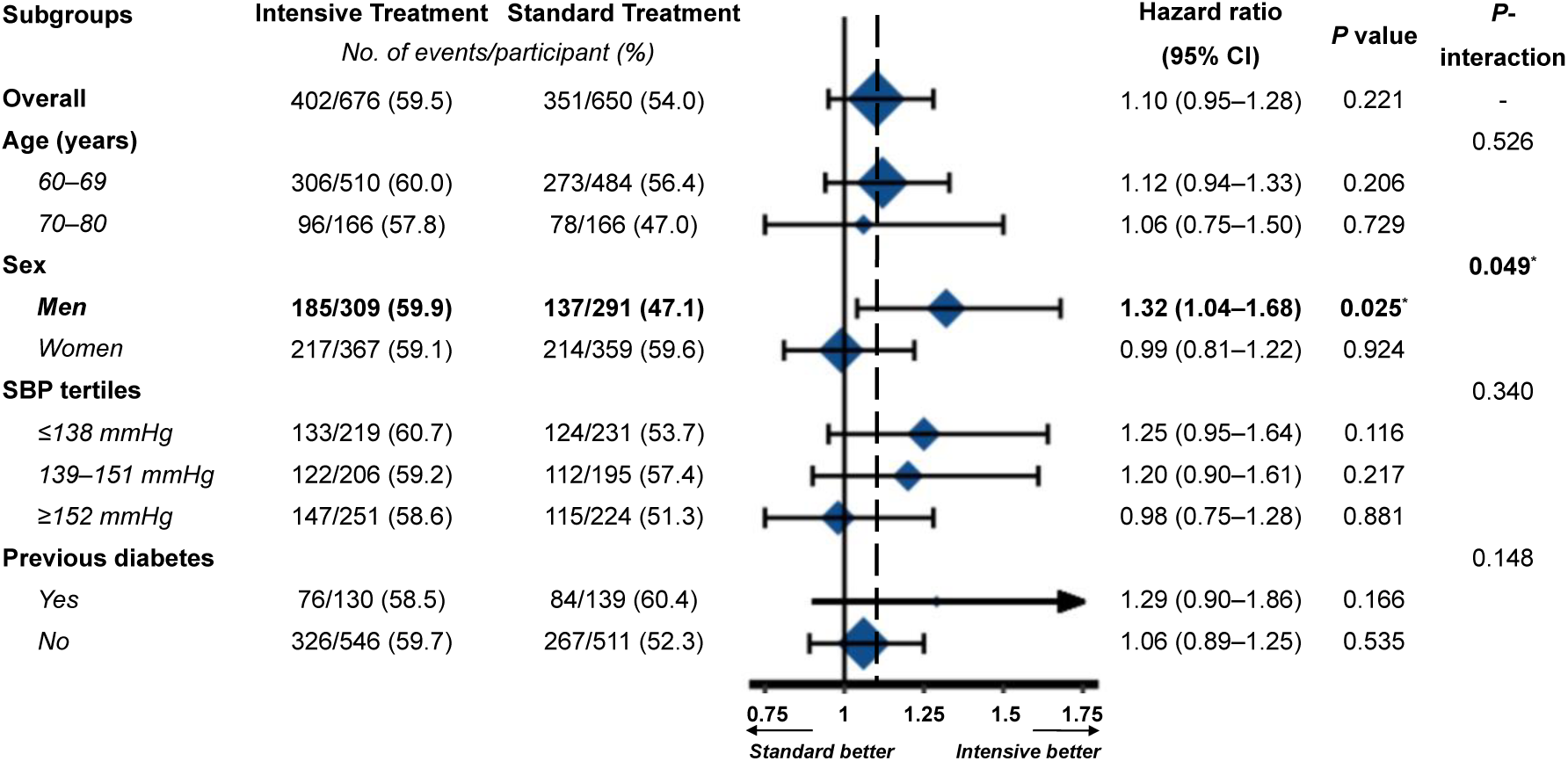
Forest plot showing the effect of intensive (versus standard) SBP-lowering treatment on LVH regression among the pre-specified subgroups during follow-up. LVH was diagnosed based on LVM/H^1.7^. The results are presented as the HR and 95% CI. The dashed line represents the overall HR. The *P*-value was calculated using Cox proportional hazards regression model. CI = confidence interval, HR = hazard ratio, H^1.7^ = height^1.7^, LVH = left ventricular hypertrophy, SBP = systolic blood pressure. N = 676 in the intensive treatment group, N = 650 in the intensive treatment group.

These results were not altered when LVH was diagnosed based on LVM indexed to H^2.7^ (Figures S4 and S5).

### LVMI regression throughout follow-up

LVMI regression occurred in both treatment groups. Random coefficient models in all patients showed that the rate of regression of LVM/H^1.7^ was greater in patients with intensive treatment than in those with standard treatment by 0.38 g/m^1.7^ per year (95% CI 0.05–0.71, *P* = 0.024) (Figure 3). The protective role of intensive SBP lowering on promoting LVMI regression was consistent among the subgroups for age, sex, SBP tertile, and diabetes mellitus status (all *P* for interaction > 0.10). These results were similar when LVM was indexed to H^2.7^ (differed by 0.23 g/m^2.7^ per year, 95% CI 0.03– 0.44, *P* = 0.025) (Figure S6). A weak but significant correlation was observed between the change in SBP and the change in LVM/H^1.7^ (*r* = 0.077, *P* = 0.000) and the change in LVM/H^2.7^ (*r* = 0.090, *P* = 0.000) throughout follow-up.

**Figure 3.**
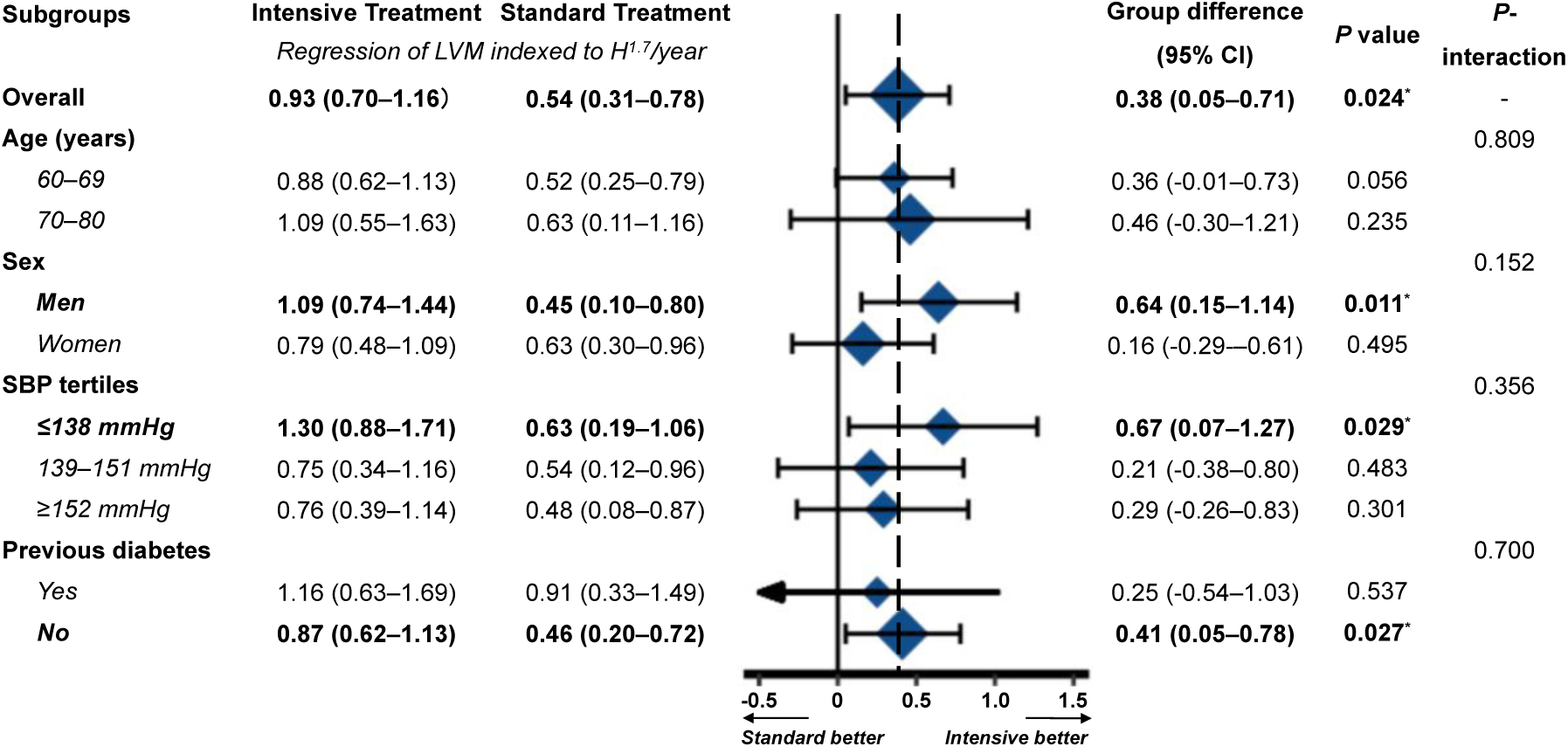
Forest plot showing the effect of intensive (versus standard) SBP-lowering treatment on LVMI regression among the pre-specified subgroups during follow-up. LVM was indexed to H^1.7^. The results are presented as the HR and 95% CI. The dashed line represents the overall HR. The *P*-value was calculated using Cox proportional hazards regression model. CI = confidence interval, HR = hazard ratio, H^1.7^ = height^1.7^, LVMI = left ventricular mass index, SBP = systolic blood pressure. N = 2915 in the intensive treatment group, N = 2794 in the intensive treatment group.

The mean annual changes in LV parameters throughout follow-up are shown in Table 2. Male patients with intensive treatment (versus standard treatment) had greater IVSTd regression (differed by 0.05 mm per year, 95% CI 0.01–0.10, *P* = 0.028) and greater PWTd regression (differed by 0.05 mm per year, 95% CI 0.01–0.10, *P* = 0.014). The changes in LVIDs, LVIDd, and LVEF were similar between the groups, regardless of sex. The changes in other echocardiographic parameters, including the diameter of the aortic sinus, left atrium, pulmonary artery, and right ventricular outflow tract; end-diastolic volume; the new development of LVEF <50%; and the new development of E/A <1, were comparable between the groups throughout follow-up, regardless of sex (Table S3).

**Table 2.**
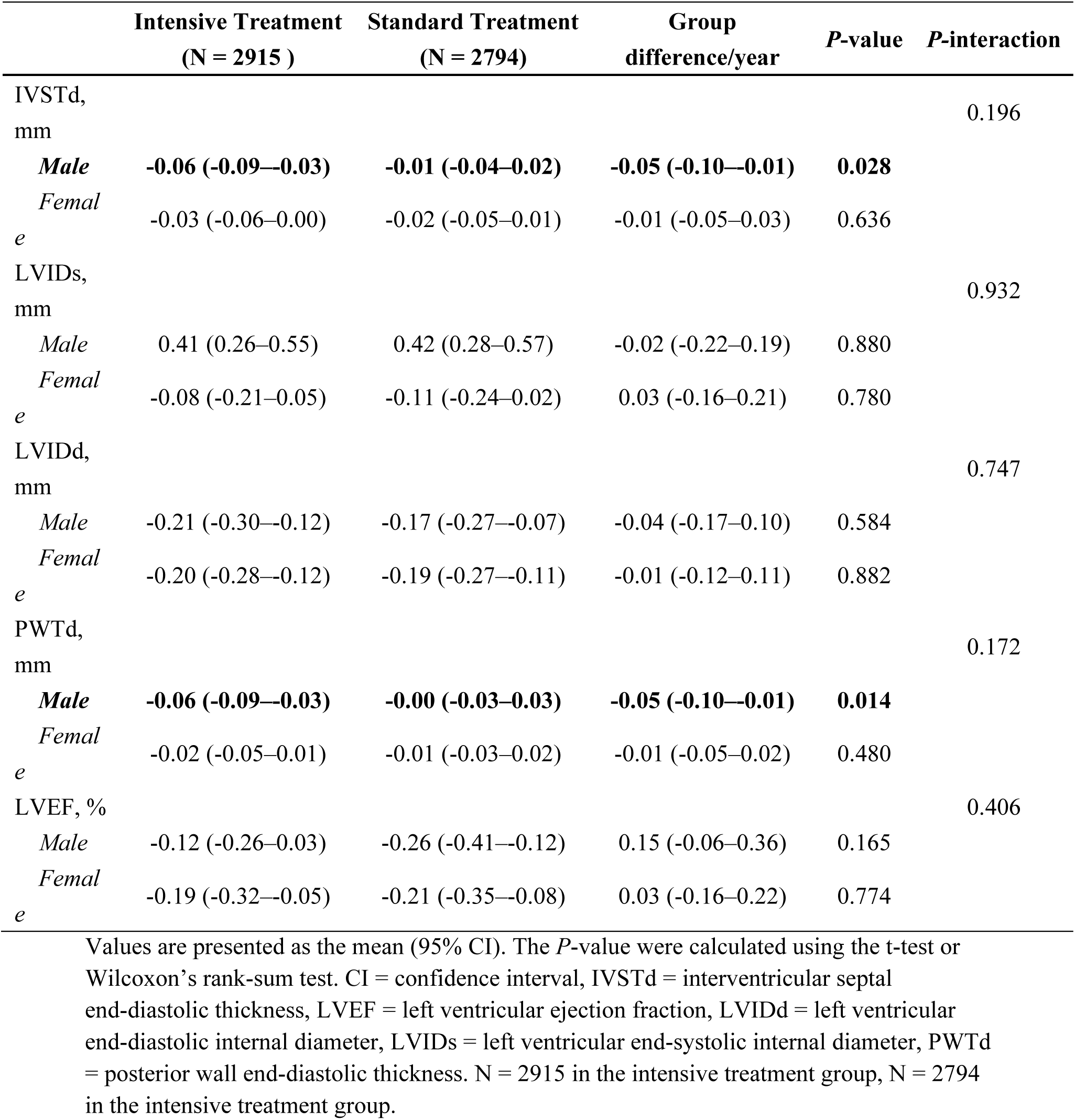
Annual changes in LV parameters throughout follow-up.

|  | Intensive Treatment<br>(N = 2915 ) | Standard Treatment<br>(N = 2794) | Group<br>difference/year | P-value | P-interaction |
| --- | --- | --- | --- | --- | --- |
| IVSTd,<br>mm |  |  |  |  | 0.196 |
| *Male* | **-0.06 (-0.09--0.03)** | **-0.01 (-0.04--0.02)** | **-0.05 (-0.10--0.01)** | **0.028** |  |
| *Femal*<br>*e* | -0.03 (-0.06--0.00) | -0.02 (-0.05--0.01) | -0.01 (-0.05--0.03) | 0.636 |  |
| LVIDs,<br>mm |  |  |  |  | 0.932 |
| *Male* | 0.41 (0.26--0.55) | 0.42 (0.28--0.57) | -0.02 (-0.22--0.19) | 0.880 |  |
| *Femal*<br>*e* | -0.08 (-0.21--0.05) | -0.11 (-0.24--0.02) | 0.03 (-0.16--0.21) | 0.780 |  |
| LVIDd,<br>mm |  |  |  |  | 0.747 |
| *Male* | -0.21 (-0.30--0.12) | -0.17 (-0.27--0.07) | -0.04 (-0.17--0.10) | 0.584 |  |
| *Femal*<br>*e* | -0.20 (-0.28--0.12) | -0.19 (-0.27--0.11) | -0.01 (-0.12--0.11) | 0.882 |  |
| PWTd,<br>mm |  |  |  |  | 0.172 |
| *Male* | **-0.06 (-0.09--0.03)** | **-0.00 (-0.03--0.03)** | **-0.05 (-0.10--0.01)** | **0.014** |  |
| *Femal*<br>*e* | -0.02 (-0.05--0.01) | -0.01 (-0.03--0.02) | -0.01 (-0.05--0.02) | 0.480 |  |
| LVEF, % |  |  |  |  | 0.406 |
| *Male* | -0.12 (-0.26--0.03) | -0.26 (-0.41--0.12) | 0.15 (-0.06--0.36) | 0.165 |  |
| *Femal*<br>*e* | -0.19 (-0.32--0.05) | -0.21 (-0.35--0.08) | 0.03 (-0.16--0.22) | 0.774 |  |

Values are presented as the mean (95% CI). The *P*-value were calculated using the t-test or Wilcoxon's rank-sum test. CI = confidence interval, IVSTd = interventricular septal end-diastolic thickness, LVEF = left ventricular ejection fraction, LVIDd = left ventricular end-diastolic internal diameter, LVIDs = left ventricular end-systolic internal diameter, PWTd = posterior wall end-diastolic thickness. N = 2915 in the intensive treatment group, N = 2794 in the intensive treatment group.

### Correlation between LVH and the primary outcome

Among the patients in the STEP trial with echocardiography data included in this analysis, a total of 203 primary composite events occurred. Patients with baseline LVH or newly developed LVH during the trial presented an increased risk of cardiovascular events (HR 1.69, 95% CI 1.20–2.40, *P* = 0.003) compared with patients without LVH. In the same model, each 1-standard deviation (16.4 g/m^1.7^) increase in mean LVM/H^1.7^ was associated with an increase in the risk of CVD events by 48% (HR 1.48, 95% CI 1.27–1.73, *P* < 0.001).

Intensive treatment was associated with a 27% lower risk (HR 0.73, 95% CI 0.54– 1.00, *P* = 0.051) of the primary outcome, which was attenuated to a 26% lower risk (HR 0.74, 95% CI 0.54–1.01, *P* = 0.056) when adjusting for LVH as a time-varying covariate, and a 25% lower risk (HR 0.75, 95% CI 0.55–1.03, *P* = 0.074) when adjusting for LVM/H^1.7^ as a time-varying covariate. The mediation analyses indicated a small but significant role for LVM/H^1.7^ (4.0%, 95% CI 0.7%–21%, *P* = 0.04) in mediating the effect of intensive SBP lowering on cardiovascular events (Table 3).

**Table 3.**
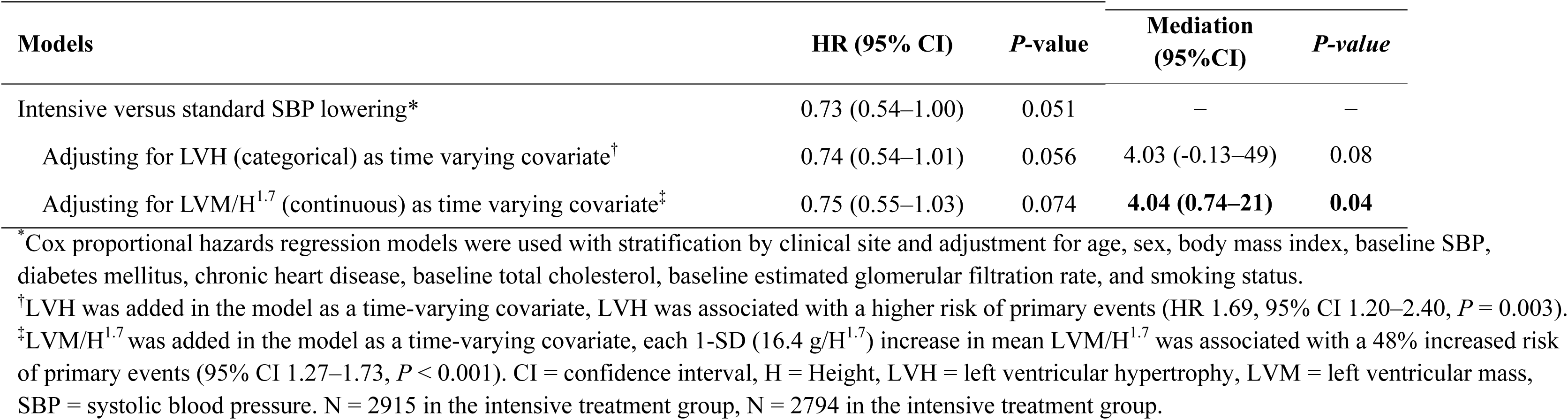
Effect of intensive SBP lowering on the primary outcome with adjustment for LVH and LVMI as time-varying covariates.

| Models | HR (95% CI) | P-value | Mediation<br>(95%CI) | P-value |
| --- | --- | --- | --- | --- |
| Intensive versus standard SBP lowering* | 0.73 (0.54–1.00) | 0.051 | – | – |
| Adjusting for LVH (categorical) as time varying covariate<sup>†</sup> | 0.74 (0.54–1.01) | 0.056 | 4.03 (-0.13–49) | 0.08 |
| Adjusting for LVM/H<sup>1.7</sup> (continuous) as time varying covariate<sup>‡</sup> | 0.75 (0.55–1.03) | 0.074 | **4.04 (0.74–21)** | **0.04** |

\*Cox proportional hazards regression models were used with stratification by clinical site and adjustment for age, sex, body mass index, baseline SBP, diabetes mellitus, chronic heart disease, baseline total cholesterol, baseline estimated glomerular filtration rate, and smoking status. <sup>†</sup>LVH was added in the model as a time-varying covariate, LVH was associated with a higher risk of primary events (HR 1.69, 95% CI 1.20–2.40, *P* = 0.003). <sup>‡</sup>LVM/H<sup>1.7</sup> was added in the model as a time-varying covariate, each 1-SD (16.4 g/H<sup>1.7</sup>) increase in mean LVM/H<sup>1.7</sup> was associated with a 48% increased risk of primary events (95% CI 1.27–1.73, *P* < 0.001). CI = confidence interval, H = Height, LVH = left ventricular hypertrophy, LVM = left ventricular mass, SBP = systolic blood pressure. N = 2915 in the intensive treatment group, N = 2794 in the intensive treatment group.

The correlation between LVH and the primary outcome was comparable in male and female patients (Tables S4 and S5). These results were similar when LVM was indexed to H^2.7^ in the sensitivity analysis (Table S6).

### Intra-rater and inter-rater reproducibility

The intraclass correlation coefficients of intra-rater variability for IVSTd, LVIDd, and PWTd were 0.96, 0.98, 0.98, respectively (all *P* < 0.001). The intraclass correlation coefficients of inter-rater variability for IVSTd, LVIDd, and PWTd were 0.95, 0.97, 0.96, respectively (all *P* < 0.001) (Figure S7).

## DISCUSSION

In this study, we evaluated the effect of intensive (versus standard) SBP lowering on echocardiographic LV geometry and its role in mediating the cardiovascular benefits associated with intensive SBP lowering. The key findings were as follows. First, compared with standard SBP lowering, intensive SBP lowering was associated with greater LVMI regression, a lower risk of LVH development, and a similar rate of LVH regression in older patients with hypertension. Second, the beneficial effect of intensive SBP lowering against LVH development and toward LVH regression was more pronounced in males than in females. Third, the favorable effect of intensive SBP lowering on LVMI regression partially explained the cardiovascular benefits associated with SBP lowering. The present study provides the first direct evidence of the favorable effect of intensive SBP lowering on LV geometry and LVH evaluated by imaging.

Consistent with previous studies where LVH was assessed by ECG, the risk towards new LVH development was significantly reduced in patients with intensive SBP lowering, whereas the rate of baseline LVH regression was similar between patients receiving intensive SBP lowering and patients receiving standard SBP lowering. These findings suggest that aggressive antihypertensive management should be instigated as soon as possible to prevent target organ damage, given that when hypertensive target organ damage is advanced, reversal may be difficult. Unexpectedly, despite similar changes in the follow-up SBP between males and females, sex differences exist in the LV geometry in response to intensive SBP lowering therapy. In patients with intensive SBP treatment, the lower risk of new LVH development and greater rate of baseline LVH regression were only observed in males. Additionally, intensive treatment was associated with regression of IVSTd and PWTd in male patients, but not in female patients. The Losartan Intervention For Endpoint reduction in hypertension (LIFE) study also observed that women had less regression of ECG-LVH than men during antihypertensive therapy(28). The reason underlying these sex differences remains unclear, one possible explanation is that adult women have significantly higher SBP augmentation indices, characterized by an increased contribution of a secondary late-systolic peak to central aortic pressure. The late-peaking systolic pressure is associated with greater LVH, even with similar SBP(29).

Another novel feature of this study was the availability of clinical prognosis data to examine whether LVM regression reduces cardiovascular risk independent of BP. In the Framingham Heart Study, regression of the ECG Cornell voltage was associated with a lower risk of cardiovascular disease, whereas progression of the Cornell voltage identified individuals at an increased risk of cardiovascular disease. Furthermore, LVH regression has been reported to reduce the incidence of cardiovascular morbidity and mortality independent of BP reduction(30). Unexpectedly, the favorable effect of intensive SBP lowering on echocardiographic LVMI regression only explains small partial of the reduction in the risk of the primary outcome, which is consistent with previous findings from the SPRINT(13), SPRINT-HEART(19), and STEP-ECG studies(15). The beneficial effect of intensive SBP lowering on cardiovascular risk reduction may occur via mechanisms other than LVM modulation. Another possible explanation is that LV geometry only mediates the effect of intensive SBP lowering on certain cardiovascular outcomes(31). The present findings do not refute the potential clinical importance of echocardiographic LV geometry in the context of intensive SBP lowering, which should continue to be considered an important target.

### Limitations and Strengths

This study has several limitations and strengths. First, the study only examined the effect of different SBP targets, and the impact of lowering SBP with specific drugs cannot be separated. Second, variability may have existed in the echocardiography measurements across sites, despite standardized observer training and use of rigorous quality control measures throughout the trial. Third, our findings may have been confounded by unmeasured factors that mediate cardiac hypertrophy (e.g., activity of the renin-angiotensin and sympathetic nervous systems, abnormalities in lipid metabolism, and inflammation)(32, 33). Finally, our findings may not be generalizable to patients with a history of stroke, those aged <60 years, and those aged >80 years, all of whom were excluded from the STEP trial.

The strengths of the study are that it is the largest RCT to date to investigate the effect of intensive SBP lowering on echocardiographic LV geometry, it included a diverse population (including patients with diabetes mellitus), the treatment groups were balanced at baseline, and the intended differences in SBP were achieved and maintained in both groups during follow-up.

### Conclusions

In older patients with hypertension without a history of stroke, LVMI regression was significantly more rapid in patients who underwent intensive SBP-lowering treatment (SBP target: 110 to <130 mmHg) than in those who underwent standard SBP-lowering treatment (SBP target: 130 to <150 mmHg). This favorable effect partially explains the reduction in cardiovascular events associated with intensive SBP lowering in the STEP trial. Our findings provide further evidence regarding the association between intensive SBP therapy and subclinical cardiac protection. From a practical perspective, the findings are in close agreement with previous reports and support the need to intensify therapeutic strategies for hypertension, which affects a large proportion of the general population worldwide.

### Perspectives

#### Clinical Competencies

In older patients with hypertension without a history of stroke, intensive SBP-lowering treatment (SBP target: 110 to <130 mmHg) is preferred than standard SBP-lowering treatment to obtain additional cardiovascular benefits in terms of echocardiographic LV geometry.

#### Translational Outlook

Although this is a relatively short-term study (median of 2.6 years), longer-term follow up of STEP will lead to better understanding of the comparative benefit by intensive SBP lowering.

## Data Availability

The data that support the findings of this study are available from the corresponding author upon reasonable request.

## Acknowledgments

We sincerely thank all members of the STEP Study Group and the study participants for their contribution to this trial. We thank Emily Woodhouse, PhD, from Liwen Bianji (Edanz) (www.liwenbianji.cn) for editing the English text of a draft of this manuscript.

## LIST OF ABBREVIATIONS

IVSTd: interventricular septal end-diastolic thickness
LVH: left ventricular hypertrophy
LVIDd: left ventricular end-diastolic internal diameter
LVMI: left ventricular mass index
PWTd: posterior wall end-diastolic thickness
RCT: randomized controlled trial
RWT: relative wall thickness
SBP: systolic blood pressure
STEP: Strategy of Blood Pressure Intervention in the Elderly Hypertensive Patients Trial

